# Attitudes of multimorbid patients to surviving future acute illness and subsequent functional disability: A systematic review

**DOI:** 10.1101/2020.06.03.20121293

**Authors:** Angela S McNelly, Luke Flower, Timothy J Stephens, AJ Fowler, Rupert M Pearse, John Prowle, Zudin A Puthucheary

**Author notes:** **Correspondence to:** Dr. Angela McNelly Critical Care and Perioperative Medicine Research Group, Adult Critical Care Unit, Royal London Hospital, London, E1 1BB, United Kingdom.

## Abstract

**BACKGROUND:** Multimorbid patients have worse outcomes following acute hospitalisation. These include increased mortality as an in-patient and after hospital discharge, and increased morbidity and dependence requiring greater use of care facilities. The literature is unclear on the views and wishes of multimorbid patients regarding the outcomes of acute hospitalisation, specifically regarding survival with additional functional disability following acute illness. This is increasingly relevant, with the recent National Institute for Health and Care Excellence (NICE) guidance on admission to hospital and critical care being based on the presence of comorbidities and function as opposed to numerical age.

**Objectives:** We performed a systematic review to assess the current qualitative literature exploring attitudes, wishes and perspectives of adult patients with multimorbidity on surviving future acute illness and subsequent acquired functional disability.

**METHODS:** *Eligibility criteria:* Eligible studies addressed the attitudes, wishes and perspectives of multimorbid adults to illness and treatment-acquired disability using qualitative methods. Information sources: A search of PubMed, Embase, and CINAHL databases was conducted from database inception through April 2020. References lists from selected papers and NICE Guidelines on Multimorbidity (NG56) were searched iteratively for additional relevant articles.

*Review methods:* Two researchers reviewed candidate full texts independently. Relevant data was extracted to an evidence table. The risk of bias was avoided by adhering to the previously published extensive search strategy and use of qualitative methodology.

**RESULTS:** From 35606 records of which 6370 were duplicates, 20 full texts were reviewed for inclusion, but none met the eligibility criteria. Coverage of domains of importance to multimorbid adults and those highlighted in the NICE guidelines on multimorbidity (NG56) by the 20 short-listed papers was determined; no publications were found to address all domains.

**DISCUSSION:** No studies were identified which have applied appropriate qualitative methodology to understand the wishes, attitudes, and preferences of multimorbid adults regarding treatment and outcomes of acute illness. Such enquiries need to be urgently undertaken to inform and progress policy and clinical practice relating to decisions around admission to hospital and critical care.

**OTHER:** *Funding:* National Institute of Health Research (NIHR) Research for Patient Benefit grant; NIHR Programme grant; NIHR Doctoral Research Fellowship.

*Registration:* PROSPERO International Prospective Register of systematic reviews (CRD: 42019155028) https://www.crd.vork.ac.uk/PROSPERO/

## Introduction

A quarter of the United Kingdom population are multimorbid, defined as the presence of two or more long-term medical conditions.^1^ This proportion increases to two-thirds of those over the age of 65,^2^ reflective of a global trend of increasing chronic disease amongst the aging population of high-income countries.^3^ Furthermore, multimorbidity is becoming increasingly prevalent in the middle-aged population.^4 5^ Consequently we expect the multimorbid population will grow, representing a global health challenge. Multimorbid patients have worse outcomes following acute hospitalisation. These include in-hospital and post-hospital rates of death, as well as greater morbidity and dependence resulting in the use of care facilities instead of being cared for at home.^6 7 8^ Attitudes to this acquired disability are ill defined regarding the consequences of acute hospitalisation - specifically in the context of survival with additional illness and acquired functional disability.

Surveys of healthy people reveal that for many, loss of independence, being a burden to family, or being admitted to a care facility are considered worse than death^9- 11^; however it is unclear whether individuals retain these attitudes overtime. The James Lind Alliance lists maintaining independent living as one of the top 10 priorities for patients with multimorbidity.^12^ Similar findings were seen in patients with chronic disease,^13^ although for some of these patients, relatively higher levels of disability were considered to be better than death, findings potentially explained by response shift phenomena (i.e. a change in an individual’s values regarding their health, in the way they perceive severity of disability, or their definition of an unacceptable level of health) and/or underestimation of adaptation (the degree to which a person learns to adjust to a new level of disability and maintain subjective quality of life). ^14 15^ However, how multimorbid patients view the trade-off between quality of life and survival post-acute illness is not known.

While literature exists on perceptions and wishes of the elderly patient, direct transposition to the multimorbid cannot be assumed when it comes to acute illness decision-making.^16^ Numerical age is considered to be less important than functional age.^17^ In the recent guidance related to acute coronavirus infections (NG159), the National Institute for Health and Care Excellence (NICE) stated that decisions to admit to critical care need to take into account the likelihood of recovery with an outcome acceptable to the patient. They further state that these decisions should be based on *“acute pathology, comorbidities and severity of illness”* as opposed to numerical age.^18^ This is in keeping with NICE guidance on multimorbidity (NG56) which also prioritises function and does not consider numerical age an important factor.^19^

Improving the care of multimorbid patients is considered a high research priority and central to this is the involvement of patients in decision making, or in the setting of an incapacitated patient, facilitating their previously expressed wishes.^20^ However, the breadth of existing research in this important area has not been well documented. Accordingly, we performed a systematic review, with a primary objective of assessing the current body of qualitative literature that explores the attitudes, wishes and perspectives of patients with multimorbidity on surviving future acute illness and subsequent acquired functional disability.

## Methods

The study protocol was registered prior to starting our review with the PROSPERO International Prospective Register of systematic reviews (CRD: 42019155028) and is reported in line with the Preferred Reporting Items for Systematic Reviews and Meta-Analyses (PRISMA) guidelines.

### Information Sources and Eligibility Criteria

We electronically searched MEDLINE, the Excerpta Medica database (EMBASE) and the Cumulative Index of Nursing and Allied Health Literature (CINAHL) database for English language original articles in peer reviewed journals, excluding conference proceedings and publications in abstract form only.

We included any study that used qualitative methods, i.e. thematic analysis of findings derived from patient interviews, narratives, observations, and reports. Eligible participants were adult patients with multimorbidity (defined as age ≥18 years and ≥2 chronic stable diseases) and did not include family members or healthcare workers.^19^ We included studies on a research question or topic of enquiry focussed on the perspectives, opinions, or perceptions of multimorbid patients on decision making concerning hospitalisation and likely outcomes.We excluded studies that collected only quantitative data (including from structured surveys and validated questionnaires) as well as reviews, commentaries, opinion-pieces, and editorials.

### Search Strategy

Three searches were undertaken to ensure all relevant publications were identified. MEDLINE, EMBASE and CINAHL were searched from the start of indexing until 28th October 2019 for the first search, to March 12^th^ 2020 for the second and to April 24^th^ 2020 for the third one, using the Healthcare Databases Advanced Search portal [https://hdas.nice.org.uk/1. Terms were combined with Boolean operators to identify studies reporting important qualitative patient outcomes amongst adults with multiple chronic illnesses. The full search strategies that were performed are listed in tables 1-3 in the Supplementary Appendix. After the first round of record screening and full text eligibility for each search, additional potentially relevant terms were extracted from short-listed papers, and further searches were performed to capture any other relevant records. In addition, snowball methods, pursuing references of references and electronic citation tracking, were used as is recommended for reviews of complex evidence.^21^

**Table 1:**
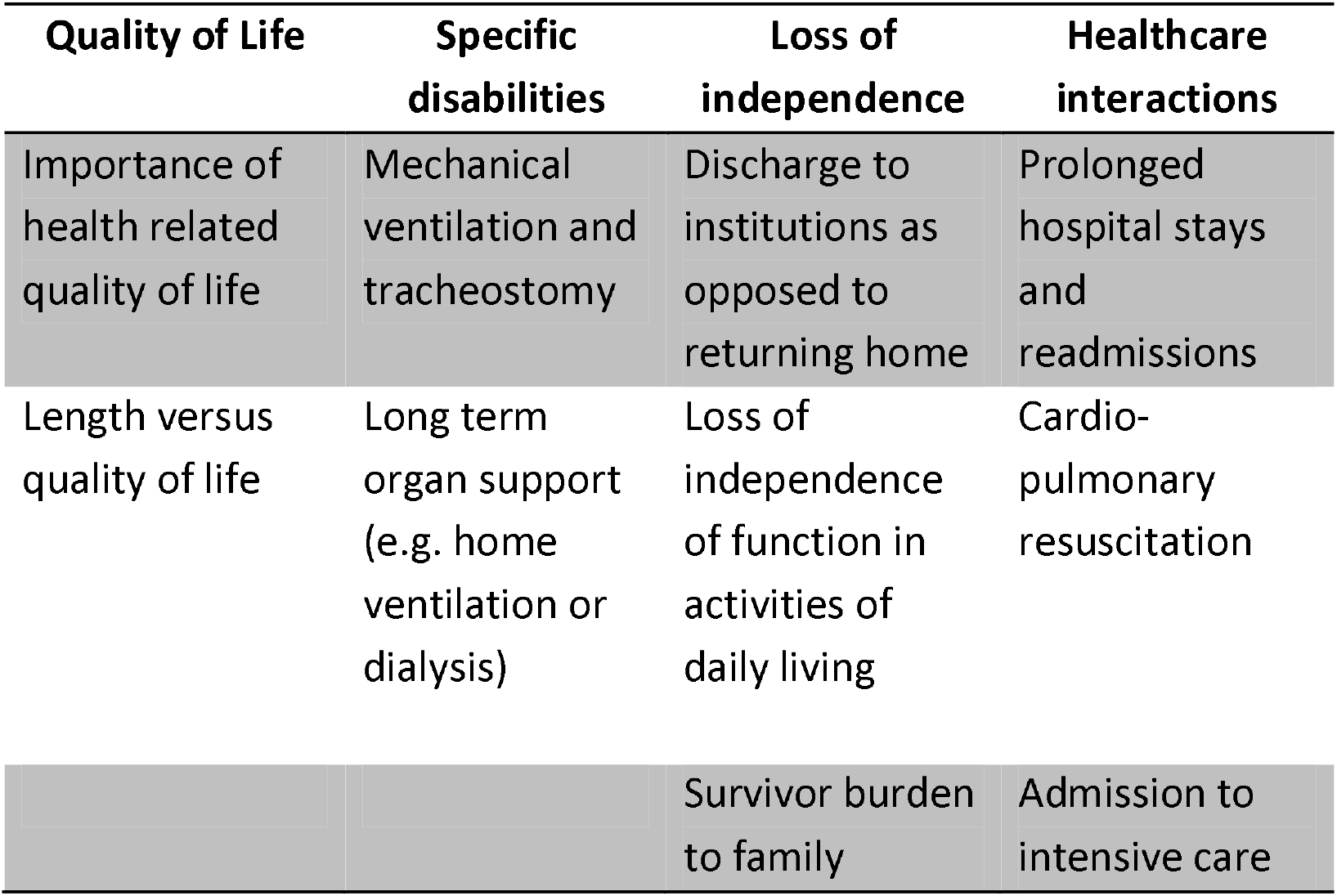
Thematic domains of importance to multimorbid adult patients in regard to consequences of surviving critical illness.

**Table 2:** Evidence of eligibility for inclusion of short-listed references. YES = Meets search criteria; NO = Does not meet search criteria.

| Author | Qualitative methods |  | Topic | Population |
| --- | --- | --- | --- | --- |
|  | Interviews | Thematic Analysis | Future Perspective of Acquired Disability | Multimorbid |
| Anthony<sup>36</sup> | YES | YES | NO | NO |
| Ohnsorge<sup>33</sup> | YES | YES | YES | NO |
| Ridgeway<sup>35</sup> | YES | YES | NO | YES |
| Lamas<sup>34</sup> | YES | YES | YES | NO |
| Ford<sup>28</sup> | NO | NO | NO | NO |
| Schoenborn<sup>27</sup> | NO | NO | NO | NO |
| Karel<sup>31</sup> | NO | NO | NO | NO |
| Raue<sup>30</sup> | NO | NO | NO | NO |
| Teno<sup>32</sup> | NO | NO | NO | YES |
| Lloyd<sup>42</sup> | YES | NO | YES | YES |
| Honeybul<sup>37</sup> | YES | YES | YES | NO |
| Karlsson<sup>38</sup> | YES | YES | NO | YES |
| Lamas<sup>25</sup> | NO | NO | YES | NO |
| Milnes<sup>26</sup> | NO | NO | YES | NO |
| Rubin<sup>13</sup> | NO | NO | YES | NO |
| Mendelsohn<sup>24</sup> | NO | NO | YES | NO |
| Gainer<sup>39</sup> | YES | YES | NO | YES |
| Thomas<sup>29</sup> | NO | NO | NO | NO |
| Scheunermann<sup>40</sup> | YES | YES | NO | YES |
| Williams<sup>41</sup> | YES | YES | NO | YES |

### Selection Process

For each search, two authors (ZAP and AM search 1; ASM and LF searches 2 and 3) independently screened records (titles and abstract). Full papers were retrieved for records with relevant abstracts and reviewed by the two researchers. Full texts of potentially eligible articles were then screened for inclusion by the two authors acting independently. Disagreements were resolved by discussion and consensus or with a third author (TJS) where consensus could not be reached.

### Data Collection

Data was extracted pertaining to (i)details of the qualitative methodology used; (ii)the specific participant cohort studied; and (iii) participants attitudes, wishes and perspectives on the topic and/or hypothetical situations presented to them for their responses. ASM and LF extracted data independently and checked whether studies met the eligibility criteria. Disagreements were resolved by discussion and consensus, with a third author (TJS) if needed. As these are qualitative studies, no protocol was required for missing data.

### Quality Assessment and Risk of Bias

It was planned that methodological quality of studies meeting search criteria would be assessed by two independent authors before inclusion in the review using the CASP tool for qualitative studies and would be included in the Supplementary Appendix.^22^ Reporting bias was minimised by adhering to the previously published extensive search strategy, and there were no competing interests to introduce bias. There is no method to assess publication bias for qualitative studies.

### Synthesis

Data analysis was planned according to established guidelines on thematic synthesis.^23^ We *a priori* stipulated that a narrative synthesis would be produced if there were few thematic data, including analysis of the domains of importance to multimorbid adults discussed in each of the short listed papers (table 1). These findings would be mapped back to the personal goals, values and priorities set out in the NICE guidance on multimorbidity and the likely outcomes (as per analysis of subgroups or subsets) for these patients were they to develop an acute illness.^19^

### Patient-Public Involvement

PPI representatives worked with us to refine the research question and are supportive of research on patient views of acquired disability post-critical illness. PPI representatives will write a plain language summary and assist with dissemination to patient groups.

## Results

Search 1 identified 8524 records, of which a short-list of 8 was extracted after removing duplicates; Search 2 identified 13366 references, of which a short list of 8 was extracted (after removing 1 duplicate from Search 1); Search 3 identified 13708 references, of which a short-list of 4 was extracted (figures 1-3). Eight additional records were identified by snowball methods, including one from the references of the NICE multimorbidity guidelines (NG56).^19^ However, when the 20 full texts were reviewed, none of the papers in any of the short-lists met the pre-specified criteria either as a result of methodological issues, patient cohort definitions or topic focuses (table 2).

**Figure 1:**
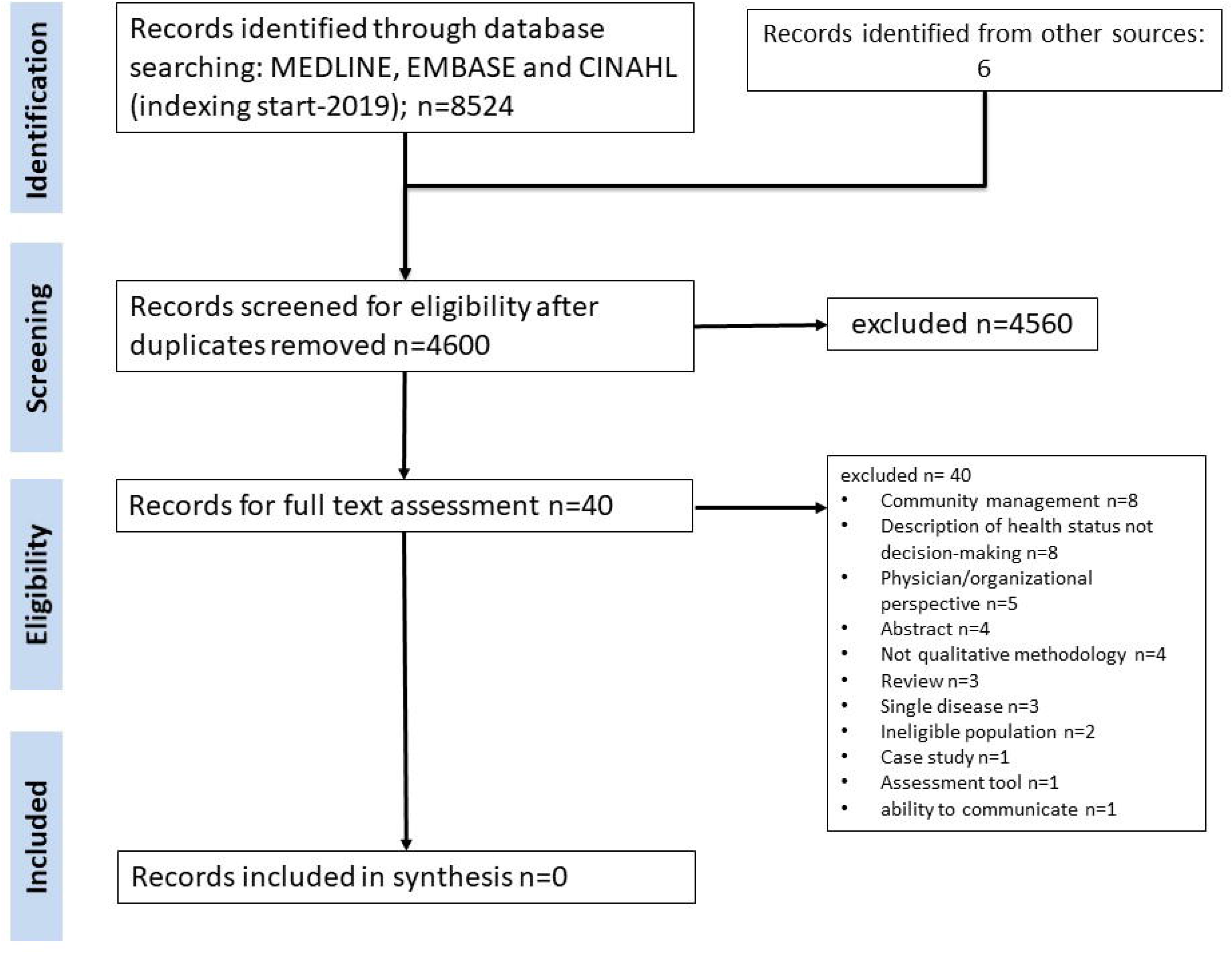
PRISMA diagram of study selection process for search 1. CINAHL=Cumulative Index to Nursing and Allied health Literature, EMBASE= Excerpta Medica Database.

**Figure 2.**
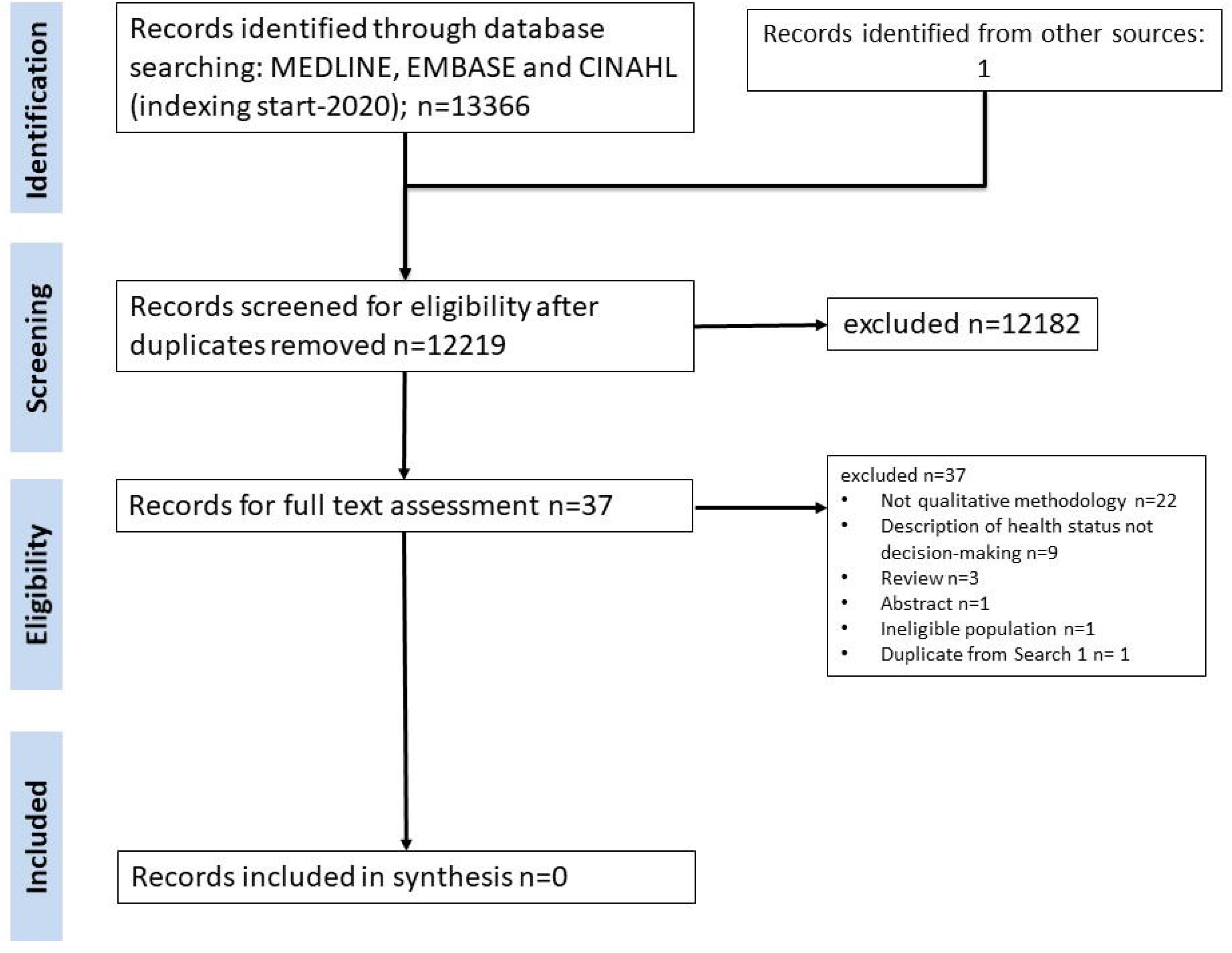
PRISMA diagram of study selection process for search 2. CINAHL=Cumulative Index to Nursing and Allied health Literature, EMBASE= Excerpta Medica Database.

**Figure 3.**
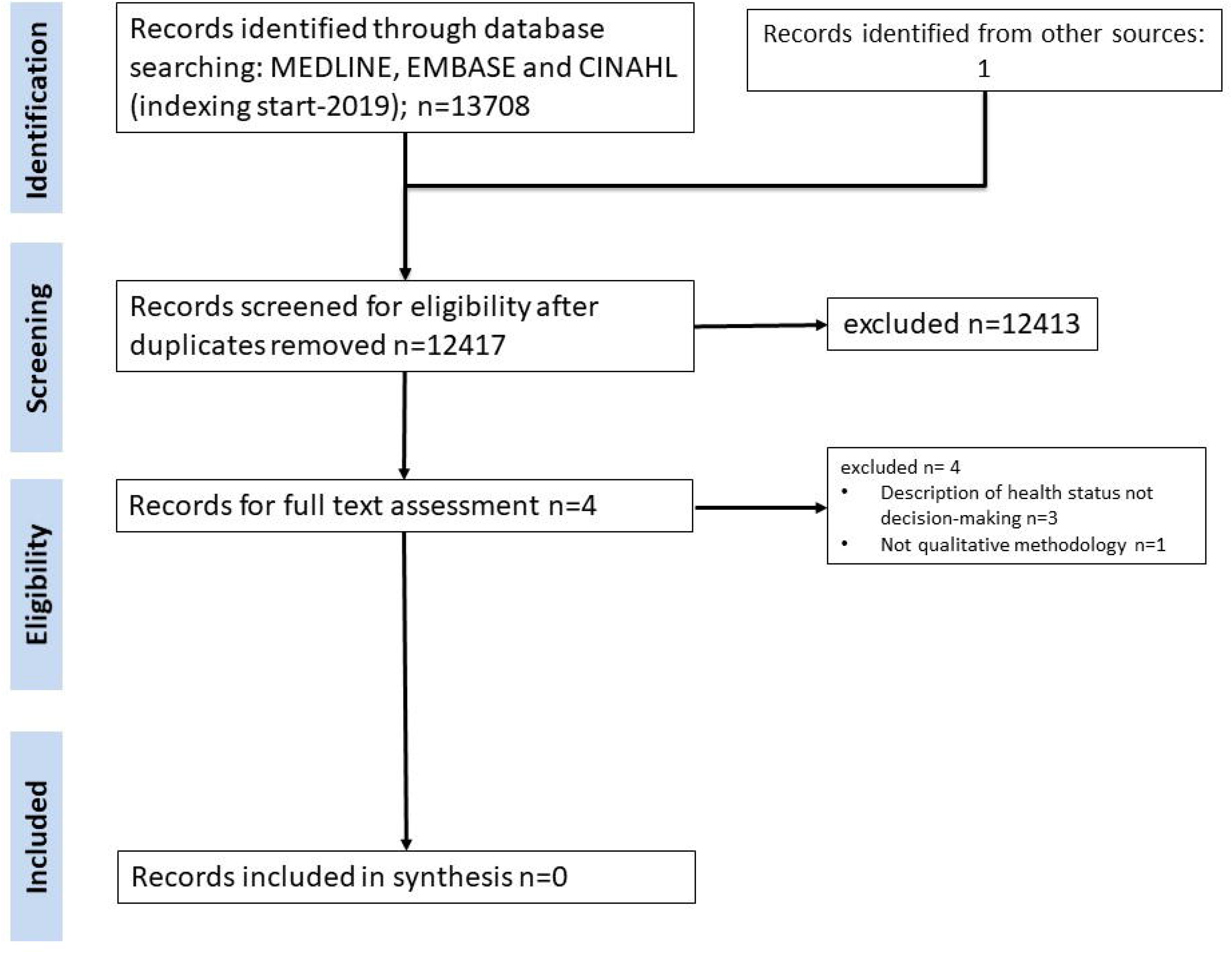
PRISMA diagram of study selection process for search 3. CINAHL=Cumulative Index to Nursing and Allied health Literature, EMBASE= Excerpta Medica Database.

### Exclusion for methodological reasons

a. <u>Surveys/auantitative analysis:</u> Ten papers used surveys or questionnaires to collect primary quantitative data from participants, which was, then subject to a subsequent quantitative analysis.^13 24-32^ Despite this, insights could be gained from some of the excluded studies: for example, the survey-based study by Rubin et al reported that hospitalised patients with multiple serious illnesses often viewed commonly encountered health states as worse than death.^13^ <u>Interviews:</u> Nine studies used structured or semi-structured interviews with thematic or Grounded theory analysis.^33-41^ One additional study did use qualitative interviews but analysed the data using time trade-off analysis.^42^ This latter study found that patients would refuse extended mechanical ventilation unless it was likely to improve prognosis, concluding that predicted Quality of Life may contribute to a similar extent to estimates of ICU survival in decision-making.

### Exclusion on basis of patient cohort

Only seven studies specifically stated that participants were multimorbid. However, several excluded studies appeared to address attitudes relevant to patients with chronic illness, and one stated that more than 90% of patients are aware of situations worse than death.^24^ Similarly, Milnes et al. concluded that patients’ priorities may be of equal or greater importance than death.^26^ Another study investigated patient expectations and experiences in a chronic critically ill population (but not specifically described as multimorbid), quoting a ventilator-dependent patient with COPD, *“It’s torture… All day like this. It’s awful”*.^34^ A study by Ohnsorge et al. focused on Wish-to-Die decisions in different hypothetical scenarios using qualitative methodology; however, this research studied an End-of-Life population, rather than those with chronic multimorbidity.^33^

### Exclusion on basis of topic focus

Only 8 of the short listed studies were considered to be on-topic.^13 24-26 33 34 37 42^ However, in each of these cases either a different population was studied or non-qualitative methods were used, meaning that none of these studies met the eligibility criteria. In four of the five qualitative studies performed in the correct patient cohort (but off-topic), the focus was on current self-management approaches,^35^ and issues around care, rather than patients’ views on acquired disability post-acute illness.^39-41^ This latter author reported that patients viewed illnesses creating discomfort as more troublesome than those that could cause death.^41^ Experiences of recovery of multimorbid individuals, rather than perspectives on future health states, were the focus for the remaining qualitative study^38^

### Mapping of data to patient goals, values and priorities

Despite the lack of papers with appropriate methodology addressing the topic in question within our defined patient cohort, some papers relevant to the topic of interest had been identified. Mapping short listed papers to subgroup domains of importance to multimorbid patients, and personal goals, values and priorities identified in the NICE guidance on multimorbidity was therefore undertaken (figure 4). Two papers met 11 of the 16 combined domains of interest,^31,33^ but no domain of interest was investigated by all papers.

**Figure 4:**
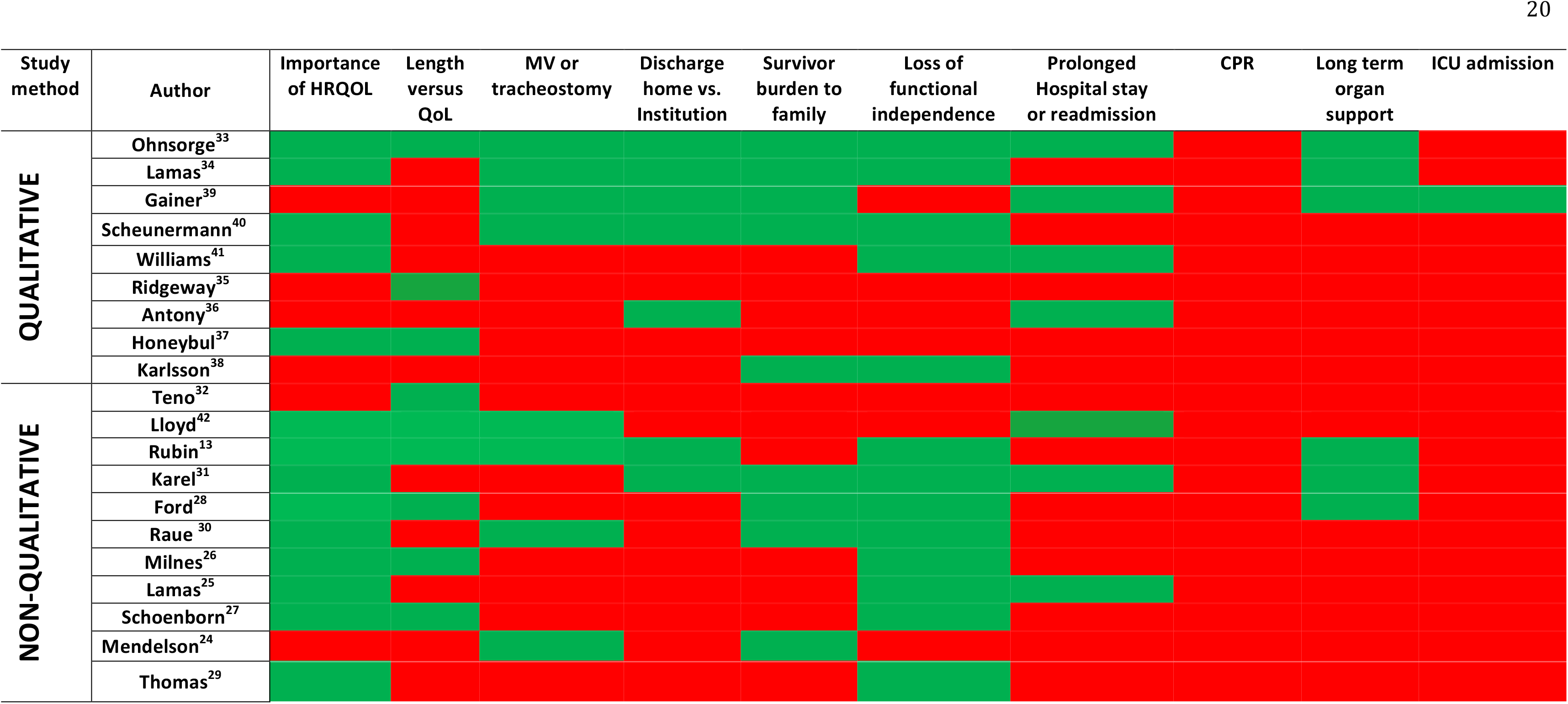
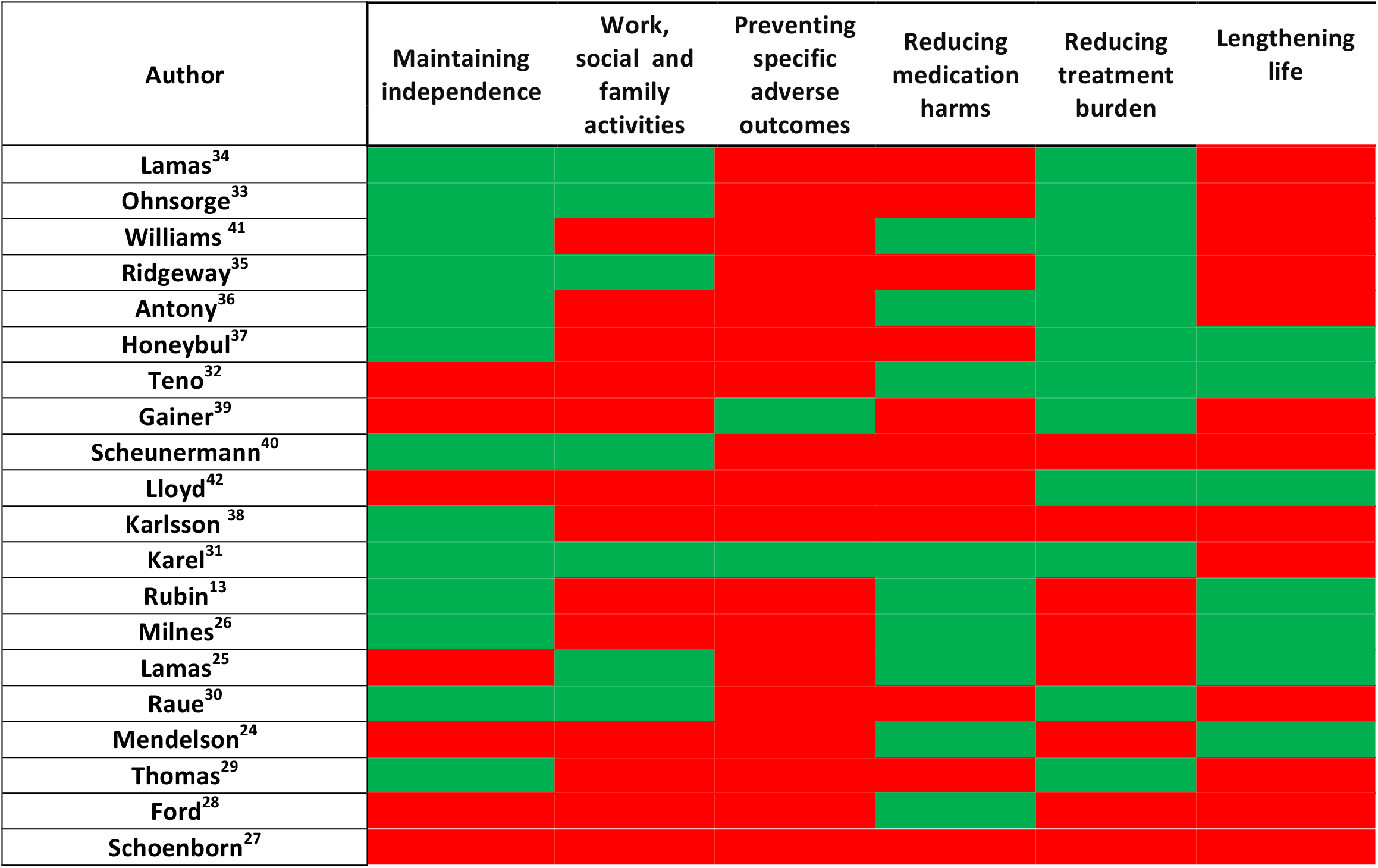
Heatmaps. I Reported views on subgroup domains of importance to multimorbid adult patients II Clarifying what’s important to people with multimorbidity from NICE Guidelines (NG56) HRQOL: Health-related quality of life; MV: Mechanical ventilation; ADL: Activities of daily living; LOS: Length of stay; ICU: Intensive care unit; CPR: Cardio-Pulmonary Resuscitation. Green: Domain included; Red: Domain not included.

## Discussion

We set out to explore the attitudes, wishes and perspectives of patients with multimorbidity on surviving future acute illness and subsequent acquired functional disability. Despite repeated searches with expanded terminology, and the iterative methods recommended for searching complex evidence, we were unable to extract a body of literature to do so. Further, the existing literature mapped poorly to the personal goals, values and priorities set out in the NICE guidance on multimorbidity, or potential outcomes resulting from surviving acute illness. Since we did not find qualitative research meeting our criteria, the planned quality assessment using the CASP tool for qualitative research was not undertaken.

We identified some striking disconnections in the coverage of domains of importance to multimorbid adult patients (figure 4): the importance of health-related quality of life and loss of functional independence were highlighted in 14 and 13 out of the 20 papers respectively; however, the need for ICU admission, a factor strongly influencing both these domains, was raised by patients in only 2 of the 20 studies. Likewise amongst the NICE parameters, 13 of the 20 reports mentioned the importance to patients of maintaining their independence; however, only 2 papers reported that preventing specific adverse outcomes (e.g. stroke) was a personal goal - despite its potential for a major and long-lasting effect on patients’ independence.

### Implications for future work

With an increasingly multimorbid population, patients need to be engaged at both policy and individual practice levels as regards preference integration into decisionmaking. To do so effectively, the onus fall on clinicians to describe alternative decisions, and elicit preferences.^43 44^ Communication is required with multimorbid patients about their views on accepting treatment options which could lead to subsequent disabilities. This needs to occur with patients directly to avoid intense emotional end-of-life decisions made by surrogates with clinicians, which can be overwhelming, impersonal and traumatic.^45^ In the acute care setting, there is evidence of poor systems of communication between older patients, their families and acute and critical care clinicians, resulting in delivery of potentially inappropriate treatments.^46-48^ Given the overlap between aging and multimorbidity, it is likely that these issues are also seen in multimorbid individuals. While some patients may not wish to discuss theoretical aspects,^27^ specific conversations on goals and values may be more acceptable.^25^ It seems important to highlight that 20% of patients go on to survive after institution of limitations in life prolonging therapies that may result in poor quality of life i.e. therapy limitations do not automatically lead to death.^49^ It may be that this is a reflection of a need for a critical care-specific measure of health related quality of life.^50^ Discussion should also highlight the findings that individuals who survive critical care support have yet to demonstrate convincing improvements in function following physical rehabilitation.^51 52^ This is likely related to loss of muscle mass,^53^ as muscle protein synthesis (the major determinant of muscle protein homeostasis and therefore muscle mass and strength) is more difficult to stimulate in the older,^54^ or inactive patient,^55^ two conditions that are increasingly common in those suffering from multimorbidity. Such patients have an attenuated response to rehabilitation^56^ and expectations therefore need to be managed appropriately.

We recommend using qualitative approaches initially as has been the focus of this systematic review. We chose to limit this review to studies that use a qualitative approach as these methodologies facilitate a detailed description, exploration and understanding of the phenomena in question, from the point of view of our population of interest.^57^ We chose to exclude quantitative questionnaire research for two reasons: the risk of bias and the lack of true patient perspective. While questionnaires do offer an objective means of collecting and reporting information about people’s perspectives, beliefs and attitudes,^58^ questionnaire research has a high potential risk of bias. This may be especially true when the topic is complex and relatively poorly understood.^59 60^ In addition, the nature of the quantitative questionnaire approach (where topics and questions are a priori defined by researchers) means the findings lack the richness and depth required if one wishes to explore and understand an issue from the perspective of those it affects.^61^

### Strengths and limitations

The strengths of this systematic review relate to the extensive searches undertaken to identify eligible papers to include in the synthesis. Two reviewers using standardised methods conducted study selection, data extraction and analysis independently. Pre-publication of our protocol on PROSEPERO ensured methodological transparency. That we did not find records eligible in methodology, population of interest and topic of discussion is a limitation of this work. We also only included abstract in the English language. We were unable to perform thematic synthesis since no studies met all the eligibility criteria, although a narrative review was performed as per the protocol.

## Conclusion

Based upon the lack of quality evidence documented in this systematic review and given the pressing need for the perspectives of multimorbid patients on treatment choices to be more clearly understood, rigorous qualitative enquiry into this complex topic is required to inform and progress policy and clinical practice.

## Data Availability

All data can be obtained from the corresponding author.

## Acknowledgements

This paper presents independent research funded by the National Institute for Health Research (NIHR). The views expressed are those of the author(s) and not necessarily those of the NIHR or the Department of Health and Social Care.

## Funding statement

ASM and ZP were supported by a National Institute of Health Research (NIHR) Research for Patient Benefit grant (PB-PG-0317-20006); AF was supported by an NIHR Doctoral Research Fellowship (DRF-2018-11-ST2-062); TS was supported an NIHR Programme grant (RP-PG-0218-20001). These researchers are independent from the funders.

All authors had full access to all data in the study and can take responsibility for the integrity of the data and the accuracy of the data analysis.

## Data sharing statement

All data can be obtained from the corresponding author.

## Contributors

ASM identified the eligibility criteria and undertook the selection process, data collection and synthesis, and drafted and revised the paper. LF identified the eligibility criteria and undertook the selection process, data collection and synthesis, and revised the draft paper. TJ identified the eligibility criteria, search strategies and undertook the selection process, data collection and synthesis, and drafted and revised the paper. AJF identified the information sources, eligibility criteria and search strategies, undertook data synthesis, and drafted and revised the paper. RMP conceptualised the study, identified the eligibility criteria and revised the paper. JP identified the information sources, eligibility criteria, search strategies, undertook data synthesis, and revised the paper. ZAP conceptualised the study, identified the eligibility criteria and undertook the selection process, data synthesis, and drafted and revised the paper. He is the guarantor.

## Mixed Competing interests

All authors have completed the ICMJE uniform disclosure form at www.icmje.org/coi_disclosure.pdf and declare: no support from any organisation for the submitted work; ASM has received non-financial support from Vitaflo ZAP has received honoraria from GlaxoSmithKline, Lyric Pharmaceuticals, Faraday Pharmaceuticals and Fresenius-Kabi; has been paid for developing and delivering educational presentations for Orion and Nestle; and has received non-financial support from Vitaflo. JP has received consultancy fees from Medibeacon Inc, Quark Pharmaceuticals Inc and Nikkiso Europe GmbH and speakers fees and/or hospitality from Baxter Inc, Nikksio Europe GmbH and Fresenius Medical Care AG. JP is an associate editor of the Clinical Kidney journal and Blood Purification, and is on the editorial board of reviewers for Intensive Care Medicine RMP has held research grants, has delivered educational presentations and/or performed consultancy work for Intersurgical, GlaxoSmithKline and Edwards Lifesciences, and holds editorial roles with the British Journal of Anaesthesia, the British Journal of Surgery and BMJ Quality and Safety; no other relationships or activities that could appear to have influenced the submitted work. LF, TJS, AJF have nothing to disclose.

## Transparency statement

The manuscript’s guarantor affirms that the manuscript is an honest, accurate, and transparent account of the study being reported; that no important aspects of the study have been omitted; and that any discrepancies from the study as planned (and, if relevant, registered) have been explained.

Ethical approval was not required for this study.

#### SUMMARY BOXES

Section 1: What is already known on this topic
- Multimorbid patients have worse outcomes following acute hospitalisation
- The NICE guidance on multimorbidity (NG56) prioritises function over numerical age in assessment and treatment planning
- NICE guideline (NG159, published March 2020) on hospital and ICU admission is based on the presence of comorbidities and function not numerical age

Section 2: What this study adds
- Our review did not identify any studies which have applied appropriate qualitative methodology to understand the attitudes and preferences of multimorbid adults regarding treatment and outcomes of acute illness
- There is an urgent need for the perspectives of multimorbid adults on treatment choices to be understood, to inform and progress policy and clinical practice relating to hospital and ICU admission

## REFERENCES

1. Yarnall AJ, Sayer AA, Clegg A, et al. New horizons in multimorbidity in older adults. Age Ageing 2017;46(6):882–88.

2. Barnett K, Mercer SW, Norbury M, et al. Epidemiology of multimorbidity and implications for health care, research, and medical education: a crosssectional study. Lancet 2012;380(9836):37–43.

3. Garin N, Koyanagi A, Chatterji S, et al. Global Multimorbidity Patterns: A CrossSectional, Population-Based, Multi-Country Study. J Gerontol A Biol Sci Med Sci 2016;71(2):205–14.

4. Kingston A, Comas-Herrera A, Jagger C, et al. Forecasting the care needs of the older population in England over the next 20 years: estimates from the Population Ageing and Care Simulation (PACSim) modelling study. Lancet Public Health 2018;3(9):e447-e55.

5. Kingston A, Robinson L, Booth H, et al. Projections of multi-morbidity in the older population in England to 2035: estimates from the Population Ageing and Care Simulation (PACSim) model. Age Ageing 2018;47(3):374–80.

6. Zekry D, Loures Valle BH, Graf C, et al. Prospective comparison of 6 comorbidity indices as predictors of 1-year post-hospital discharge institutionalization, readmission, and mortality in elderly individuals. J Am Med Dir Assoc 2012;13(3):272–8.

7. Zador Z, Landry A, Cusimano MD, et al. Multimorbidity states associated with higher mortality rates in organ dysfunction and sepsis: a data-driven analysis in critical care. Crit Care 2019;23(1):247.

8. Szakmany T, Walters AM, Pugh R, et al. Risk Factors for 1-Year Mortality and Hospital Utilization Patterns in Critical Care Survivors: A Retrospective, Observational, Population-Based Data Linkage Study. Crit Care Med 2019;47(1):15-22.

9. Ditto PH, Druley JA, Moore KA, et al. Fates worse than death: the role of valued life activities in health-state evaluations. Health Psychol 1996;15(5):332–43.

10. Fried TR, Van Ness PH, Byers AL, et al. Changes in preferences for life-sustaining treatment among older persons with advanced illness. J Gen Intern Med 2007;22(4):495–501.

11. Patrick DL, Pearlman RA, Starks HE, et al. Validation of preferences for life-sustaining treatment: implications for advance care planning. Ann Intern Med 1997;127(7):509–17.

12. Alliance JL. Secondary. http://www.ila.nihr.ac.uk/priority-setting-partnerships/health-with-multiple-conditions-in-old-age/downloads/Multiple-Conditions-in-Later-Life-PSP-Top-10-report.pdf.

13. Rubin EB, Buehler AE, Halpern SD. States Worse Than Death Among Hospitalized Patients With Serious Illnesses. JAMA Intern Med 2016;176(10):1557–59.

14. Sprangers MA, Schwartz CE. Integrating response shift into health-related quality of life research: a theoretical model. Soc Sci Med 1999;48(11):1507–15.

15. Ubel PA, Loewenstein G, Schwarz N, et al. Misimagining the unimaginable: the disability paradox and health care decision making. Health Psychol 2005;24(4S):S57-62.

16. van Oppen JD, Keillor L, Mitchell A, et al. What older people want from emergency care: a systematic review. Emerg Med J 2019;36(12):754–61.

17. Pollock RD, Carter S, Velloso CP, et al. An investigation into the relationship between age and physiological function in highly active older adults. The Journal of physiology 2015;593(3):657–80; discussion 80.

18. NICE. COVID-19 rapid guideline: critical care in adults. Secondary COVID-19 rapid guideline: critical care in adults 2020. https://www.nice.ore.uk/euidance/NG159.

19. NICE. Multimorbidity:clinical assessment and managment. Secondary Multimorbidityxlinical assessment and management 2016. https://www.nice.ore.uk/euidance/ne56/chapter/Recommendations.

20. Sciences AoM. Multimorbidity: a priority for global health research. Secondary Multimorbidity: a priority for global health research 2018. https://acmedsci.ac.uk/policv/policv-proiects/multimorbiditv.

21. Greenhalgh T, Peacock R. Effectiveness and efficiency of search methods in systematic reviews of complex evidence: audit of primary sources. BMJ 2005;331(7524): 1064-5.

22. Programme CAS. Secondary. https://casp-uk.net/wp-content/uploads/2018/01/CASP-Qualitative-Checklist-2018.pdf.

23. Thomas J, Harden A. Methods for the thematic synthesis of qualitative research in systematic reviews. BMC Med Res Methodol 2008;8:45.

24. Mendelsohn AB, Belle SH, Fischhoff B, et al. How patients feel about prolonged mechanical ventilation 1 year later. Crit Care Med 2002;30(7):1439–45.

25. Lamas DJ, Owens RL, Nace RN, et al. Conversations About Goals and Values Are Feasible and Acceptable in Long-Term Acute Care Hospitals: A Pilot Study. J Palliat Med 2017;20(7):710–15.

26. Milnes S, Corke C, Orford NR, et al. Patient values informing medical treatment: a pilot community and advance care planning survey. BMJ Support Palliat Care 2019;9(3):e23.

27. Schoenborn NL, Janssen EM, Boyd C, et al. Older Adults’ Preferences for Discussing Long-Term Life Expectancy: Results From a National Survey. Ann Fam Med 2018;16(6):530–37.

28. Ford D, Zapka J, Gebregziabher M, et al. Factors associated with illness perception among critically ill patients and surrogates. Chest 2010;138(1):59-67.

29. Thomas CM, Sklar MC, Su J, et al. Evaluation of Older Age and Frailty as Factors Associated With Depression and Postoperative Decision Regret in Patients Undergoing Major Head and Neck Surgery. JAMA Otolaryngol Head Neck Surg 2019.

30. Raue PJ, Morales KH, Post EP, et al. The wish to die and 5-year mortality in elderly primary care patients. Am J Geriatr Psychiatry 2010;18(4):341–50.

31. Karel MJ, Mulligan EA, Walder A, et al. Valued life abilities among veteran cancer survivors. Health Expect 2016;19(3):679–90.

32. Teno JM, Fisher E, Hamel MB, et al. Decision-making and outcomes of prolonged ICU stays in seriously ill patients. J Am Geriatr Soc 2000;48(S1):S70-4.

33. Ohnsorge K, Rehmann-Sutter C, Streeck N, et al. Wishes to die at the end of life and subjective experience of four different typical dying trajectories. A qualitative interview study. PLoS One 2019;14(1):e0210784.

34. Lamas DJ, Owens RL, Nace RN, et al. Opening the Door: The Experience of Chronic Critical Illness in a Long-Term Acute Care Hospital. Crit Care Med 2017;45(4):e357-e62.

35. Ridgeway JL, Egginton JS, Tiedje K, et al. Factors that lessen the burden of treatment in complex patients with chronic conditions: a qualitative study. Patient Prefer Adherence 2014;8:339-51.

36. Antony SM, Grau LE, Brienza RS. Qualitative study of perspectives concerning recent rehospitalisations among a high-risk cohort of veteran patients in Connecticut, USA. BMJ Open 2018;8(6):e018200.

37. Honeybul S, Gillett GR, Ho KM, et al. Long-term survival with unfavourable outcome: a qualitative and ethical analysis. J Med Ethics 2015;41(12):963–9.

38. Karlsson V, Bergbom I, Ringdal M, et al. After discharge home: a qualitative analysis of older ICU patients’ experiences and care needs. Scand J Caring Sci 2016;30(4):749–56.

39. Gainer RA, Curran J, Buth KJ, et al. Toward Optimal Decision Making among Vulnerable Patients Referred for Cardiac Surgery: A Qualitative Analysis of Patient and Provider Perspectives. Med Decis Making 2017;37(5):600–10.

40. Scheunemann LP, White JS, Prinjha S, et al. Post-Intensive Care Unit Care. A Qualitative Analysis of Patient Priorities and Implications for Redesign. Ann Am Thorac Soc 2020;17(2):221–28.

41. Williams A. Patients with comorbidities: perceptions of acute care services. J Adv Nurs 2004;46(1):13-22.

42. Lloyd CB, Nietert PJ, Silvestri GA. Intensive care decision making in the seriously ill and elderly. Crit Care Med 2004;32(3):649–54.

43. Elwyn G, Lloyd A, May C, et al. Collaborative deliberation: a model for patient care. Patient Educ Couns 2014;97(2):158–64.

44. Elwyn G, Frosch DL, Kobrin S. Implementing shared decision-making: consider all the consequences. Implement Sci 2016;11:114.

45. Cox CE, Hua M, Casarett D. A Measured Dose of Optimism for the Evolution of ICU-Based Palliative Care. JAMA 2019.

46. Heyland D, Cook D, Bagshaw SM, et al. The Very Elderly Admitted to ICU: A Quality Finish? Crit Care Med 2015;43(7):1352–60.

47. Heyland DK, Garland A, Bagshaw SM, et al. Recovery after critical illness in patients aged 80 years or older: a multi-center prospective observational cohort study. Intensive Care Med 2015;41(ll):1911-20.

48. Heyland DK, Barwich D, Pichora D, et al. Failure to engage hospitalized elderly patients and their families in advance care planning. JAMA Intern Med 2013;173(9):778–87.

49. Sprung CL, Ricou B, Hartog CS, et al. Changes in End-of-Life Practices in European Intensive Care Units From 1999 to 2016. JAMA 2019:1-12.

50. Lim WC, Black N, Lamping D, et al. Conceptualizing and measuring health-related quality of life in critical care. J Crit Care 2016;31(1):183-93.

51. Walsh TS, Salisbury LG, Merriweather JL, et al. Increased Hospital-Based Physical Rehabilitation and Information Provision After Intensive Care Unit Discharge. The RECOVER Randomized Clinical Trial. JAMA Intern Med 2015;175(6):901–10.

52. Denehy L, Skinner EH, Edbrooke L, et al. Exercise rehabilitation for patients with critical illness: a randomized controlled trial with 12 months of follow-up. Critical care (London, England) 2013;17(4):R156.

53. Puthucheary ZA, Rawal J, McPhail M, et al. Acute skeletal muscle wasting in critical illness. JAMA 2013;310(15):1591–600.

54. Kumar V, Selby A, Rankin D, et al. Age-related differences in the doseâ€”response relationship of muscle protein synthesis to resistance exercise in young and old men. The Journal of physiology 2009;587(1):211–17.

55. Phillips SM, Glover EI, Rennie MJ. Alterations of protein turnover underlying disuse atrophy in human skeletal muscle. J Appl Physiol 2009;107(3):645–54.

56. Puthucheary ZA, Denehy L. Exercise Interventions in Critical Illness Survivors: Understanding Inclusion and Stratification Criteria. American journal of respiratory and critical care medicine 2015;191(12):1464–7.

57. Ritchie J, Lewis, J. McNaughton Nicholls, C., Ormston, R. Qualitative Research Practice: A Guide for Social Science Students and Researchers: SAGE publication Ltd, 2013.

58. Gillham B. Developing a questionnaire. London: Bloomsbury, 2000.

59. Boynton PM. Administering, analysing, and reporting your questionnaire. BMJ 2004;328(7452): 1372-5.

60. Choi BC, Pak AW. A catalog of biases in questionnaires. Prev Chronic Dis 2005;2(1):A13.

61. Boynton PM, Greenhalgh T. Selecting, designing, and developing your questionnaire. BMJ 2004;328(7451):1312–5.

